# The politics and challenges of measuring mortality for humanitarian decision-making in Somalia, the Central African Republic and Bangladesh

**DOI:** 10.1101/2025.06.19.25329924

**Authors:** Jennifer Palmer, Mervat Alhaffar, Bashiru Garba, Jihaan Hassan, Ruwan Ratnayake, Andrew Seal, Mohamed Jelle, Francesco Checchi

## Abstract

Excess mortality is a fundamental metric of humanitarian crisis severity and should guide aid prioritisation. Despite intense political interest in mortality measurements, few studies have examined the political and institutional contexts in which mortality data are produced and used. To explore the challenges and opportunities of using evidence for humanitarian decision-making, we examined the political and organisational circumstances surrounding how mortality data are measured and used in three crisis contexts: Somalia, Central African Republic (CAR), and Cox’s Bazar in Bangladesh. We used a political economy approach to analyse information from 23 national level interviews, document review, and observations at global mortality estimation meetings. Insecurity and bureaucratic obstacles to measuring mortality in areas outside government control created data coverage gaps in CAR and Somalia. Combined with the effects of censorship in CAR, these gaps likely limit humanitarian actors’ understanding of interventions that might be needed. Across settings, new crises brought additional financing and innovations in measurement, though they could not always be sustained. United Nations coordination systems and programmes provided the backbone for regular measurement work and new initiatives, but data sharing challenges created inefficiencies. In Somalia and CAR, mortality measurement needs to be scaled and better coordinated to influence decisions. In the context of widespread aid cuts, crisis-affected governments may take increasing ownership of certain areas of mortality measurement work, but the political nature of mortality data means that independent actors and funding are needed to ensure data are collected, counter censorship, and build capacities of diverse stakeholders.

## 1. Introduction

Estimating mortality associated with humanitarian crises is critical to objectively assessing needs and key for guiding and evaluating humanitarian responses (Checchi, 2023). Excess mortality is arguably the most important metric of the severity of a crisis, and should inform all aid decisions affecting physical health (UNHCR, 2023). Mortality data can also be used to document violence, establish causal links with violations of international humanitarian law, and for purposes of memorialisation and restorative justice (Checchi and Roberts, 2008; Ratnayake et al., 2009). Mortality measurement could take on increasing importance in the humanitarian sector in the context of major cuts to humanitarian funding and calls for greater aid efficiency in 2025’s “humanitarian reset” (OCHA, 2025).

Mortality assessment is frequently conducted using retrospective surveys or surveillance methodologies (Checchi, 2018), but a variety of challenges impede its more systematic application (Spiegel and Robinson, 2010). A lack of vital registration systems and fast-moving dynamics within crises means that mortality data usually need to be proactively and frequently collected. Mortality measurement work is rarely accurate or comprehensive because of difficulties accessing populations, with the hardest to reach often the most vulnerable.

There is also widespread recognition of how political mortality measurement can be (Checchi and Roberts, 2008), but few studies have examined the political and institutional contexts in which mortality data are produced and used to make decisions. An important exception comes from the literature on famine politics and the Integrated Food Security Phase Classification (IPC) system.

Through a series of case studies, Maxwell and Hailey describe how infrequently mortality data are available, describing them as “the most politically charged kind” (Maxwell and Hailey, 2021, p. 8). Common political influences on the measurement of mortality and malnutrition include intimidation of field teams, real and imagined security obstacles and bureaucratic control on implementation and dissemination. The pressures on government-humanitarian consensus processes can lead to politically negotiated “Goldilocks solutions” that are “just right” because they don’t faithfully reflect evidence but all parties can live with them (page 20). There is also growing journalistic attention to exposing the ways governments at war may block and falsify the flow of data to the IPC or suppress its findings, with harmful impacts on aid and vulnerable populations, such as in Myanmar, Sudan and Gaza (Farley et al., 2024; Masri et al., 2024; McPherson et al., 2024). Academics implementing mortality surveys have also spoken out about behind-the-scenes reactions to their findings by diverse actors in Iraq, Uganda, Rwanda, Democratic Republic of the Congo, Central African Republic (CAR) and elsewhere (Checchi and Roberts, 2008; Checchi, Francesco, 2007; Maxmen, 2024).

We aimed to examine the political, operational and bureaucratic challenges that influence how mortality is measured and used for humanitarian decision-making by the range of actors present in three crisis-affected contexts: Somalia, CAR, and Cox’s Bazar in Bangladesh. Across these settings, humanitarian needs are high. The structures which have evolved to coordinate humanitarian work in each crisis setting, however, are different and enable actors to exert power in different ways. By tracing how measurement and analysis practices have emerged to produce, mobilise or limit mortality information within diverse contexts, we hope to inform ongoing efforts in the humanitarian sector to improve the use of evidence for decision-making.

## 2. Methods

### 2.1. Study design

We undertook qualitative case studies to compare system dynamics of mortality data collection and use. Our aim was to provide examples to understand common system-level challenges that need to be addressed and to identify operational and governance-related solutions that could help design a new global mechanism to encourage systematic mortality evidence generation and uptake in future crises (authors, forthcoming). We chose our case study settings because they were affected by large humanitarian crises; sufficient mortality measurement activity had taken place to generate lessons; they represented a combination of places where members of our research team had and had not worked before; and it was feasible to collect data.

### 2.2. Study settings

In the last three decades, Somalia has seen cyclical droughts, floods and acute food insecurity against a backdrop of persistent conflict. Considered a ‘failed state’ in the 1990s because of a lack of a central authority, Somalia is now a republic composed of the Federal Government and five Federal Member States. The non-state armed actor, Al-Shabaab controls large areas of rural, southern, Somalia. Because of its proneness to food insecurity and nutrition crises, Somalia has been at the centre of global nutrition and mortality measurement innovation. Somalia hosts refugees, mainly from Ethiopia and Yemen, and has a consistently large internally displaced population (IDP) including 1.2 million in privately-run IDP camps around Mogadishu alone (Adam et al., 2025).

CAR has been in civil war since 2012, with a recent peak of violence in 2021-22 when the government retook substantial territory from non-state actors in the north and west. Like other former Francophone African countries, the last decade has seen CAR’s government reject French and UN peacekeeping in favour of interventions from Russia, in what has been described as a new Cold War battleground “where news and fake news are constantly battling for supremacy” (Jackson, 2024). One in five Central Africans are internally displaced or living outside the country; CAR also hosts refugees, mainly from Sudan and Chad.

The Rohingya people, a persecuted ethnic minority from Myanmar, have experienced several large-scale forced displacements over five decades. The largest was into Bangladesh in August 2017. One million refugees mainly live in 33 overcrowded camps within Cox’s Bazar district. They are dependent on humanitarian aid, as refugees in Bangladesh face movement restrictions and are not legally permitted to work.

Both Somalia and CAR have ‘cluster’ coordination systems overseen by the UN’s Office for the Coordination of Humanitarian Affairs (OCHA). In Cox’s Bazar, multi-sector coordination within the humanitarian response is overseen by UNHCR and the Government of Bangladesh. Our institutions (e4c, SIMAD University, and LSHTM) have been key actors in mortality estimation in Somalia but not the other settings.

### 2.3. Data collection

Data collection happened between May 2024 and March 2025, and consisted of 6-10 interviews with informants in each case study context, review of mortality survey and surveillance reports from each context, observations from two global-level meetings discussing crisis-related mortality measurement and three feedback meetings convened with global stakeholders in early 2025. In the non-public meetings, we announced we were using the meetings to inform our study and followed up with some participants for further information.

Data collection was iterative, with the meetings, document review and interviews, and our own knowledge of mortality work, each contributing information to the case studies and identification of new informants for interviews. Online searches of peer reviewed literature, ReliefWeb and institutional websites were used to scope the range of mortality assessments in each setting.

Informants were from donors, UN agencies, local and international non-governmental organizations (NGOs), academic institutions and government technical agencies (Table 1). Informants were selected because they had been involved in at least one mortality measurement initiative or played a coordinating role to translate mortality evidence into humanitarian decision-making. More than one person per organisation was interviewed if they could offer different perspectives or detail. We did not receive responses from around one-third of people we approached for an interview, including from people who mentioned they would need to request permission from their organisational hierarchies because of the sensitivity of mortality measurement. Interviews about CAR and Cox’s Bazaar were conducted remotely and those for Somalia were in-person or remote.

**Table 1.** Informants consulted about each case study context, by type of organisation and gender of informant.

|  | **Somalia** | **CAR** | **Cox's Bazar** | **Total** |
| --- | --- | --- | --- | --- |
| Government of case study country | 0 | 0 | 0 | 0 |
| Foreign donor | 1 | 0 | 0 | 1 |
| UN agency | 0 | 2 | 5 | 7 |
| NGO | 3 | 6 | 0 | 9 |
| Foreign academic or technical agency | 2 | 2 | 2 | 6 |
| **Total** | **6** | **10\*** | **7** | **23** |
| # who were men | 6 | 6 | 4 | 16 |

\*Includes 2 people who provided only written answers to emailed questions (no verbal interview).

Interviews were conducted in English, followed a flexible topic guide, were audio-recorded and transcribed; two people provided written responses to emailed questions only. Further details of our approach to data collection and analysis are available in online Appendix A.

### 2.4. Data analysis

Our analysis was inspired by principles of applied political-economy analysis. Such an approach seeks to understand what is really ‘going on’ in a situation and considers how different actors’ behaviours may be facilitated or constrained by: structural and contextual factors; formal or informal norms, practices and beliefs; actors’ power and capacities; and actors’ interests and priorities (Whaites, 2017). We developed a common analysis template to summarise information about each context which we also use to present our data here: the range and evolution of mortality measurement initiatives undertaken by actors in the setting; key political, operational and bureaucratic constraints and solutions; and examples of mortality measurements influencing humanitarian decision-making. We developed timelines to track how innovations emerged (or were suppressed) in the context of political and humanitarian events and other mortality measurement initiatives (Figure 1, Figure 2, Figure 3). Interviews from Somalia, CAR and Cox’s Bazar are reported using the codes SM, CR and CB, respectively.

**Figure 1.**
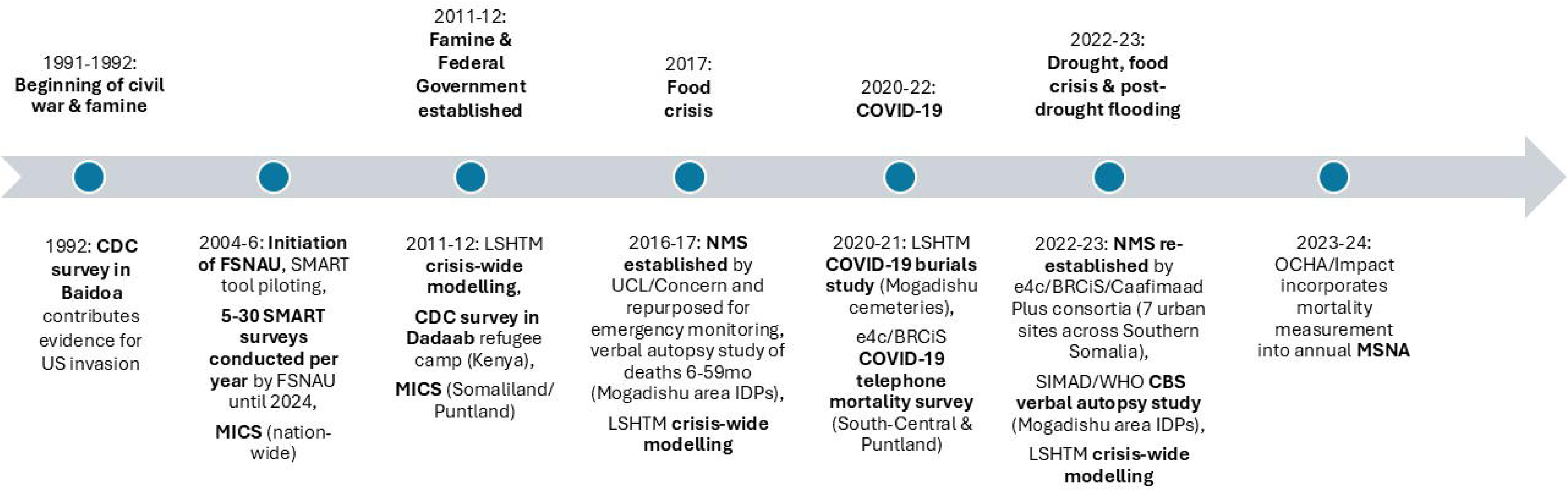
Timeline of crisis (top) and mortality measurement events (bottom) in Somalia.

**Figure 2.**
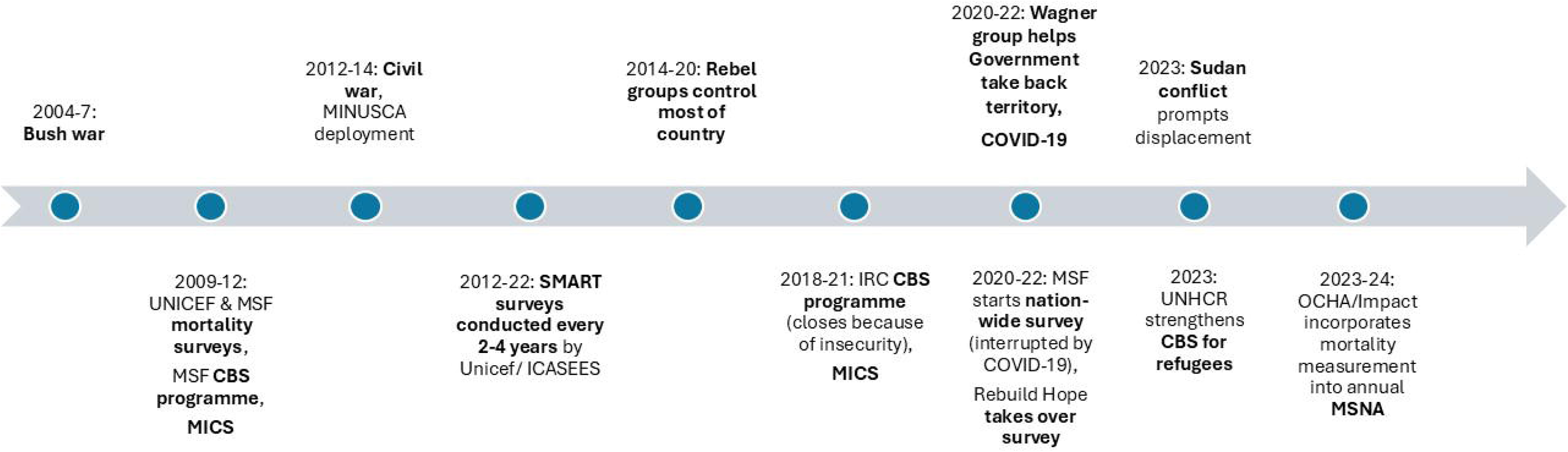
Timeline of crisis (top) and mortality measurement events (bottom) in the Central African Republic.

**Figure 3.**
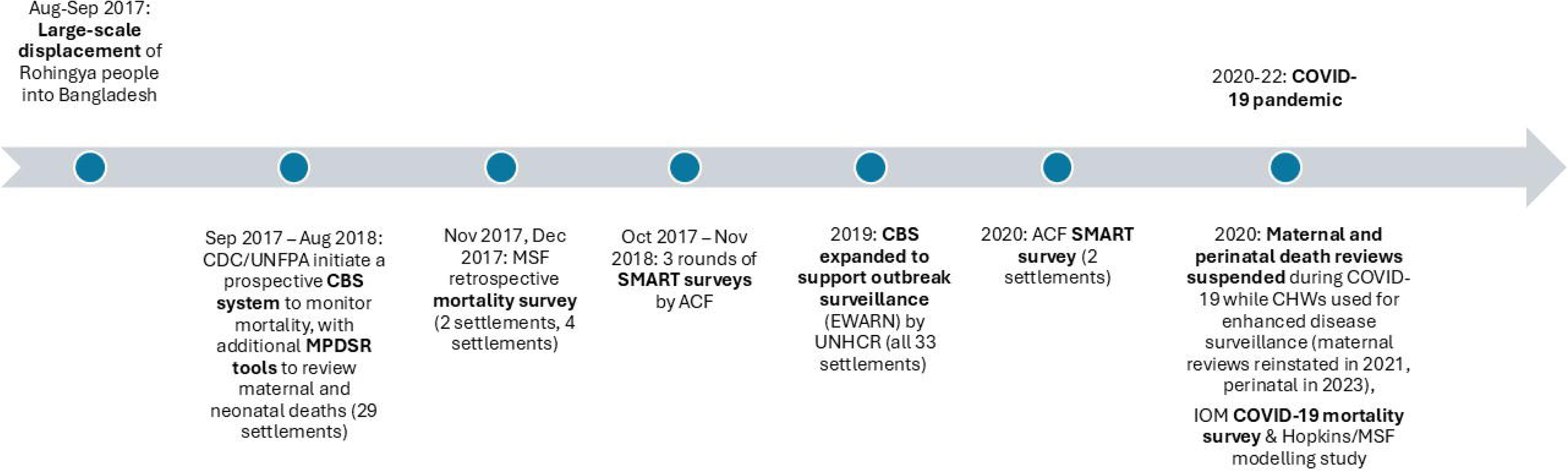
Timeline of crisis (top) and mortality measurement events (bottom) in Cox’s Bazar refugee camps, Bangladesh.

### 2.5. Ethics

This study was approved by the Research Ethics Review Committees of the London School of Hygiene and Tropical Medicine in the UK (ref: 31136) and SIMAD University in Somalia (ref: 2024/SU-IRB/FMHS/P0012). All informants provided written consent for use of their contributions shared in interviews, via email, and in stakeholder meetings. Some material shared ‘off the record’ has been excluded. Before external presentations and publishing written findings, we asked informants permission to use quotes and verified that we had accurately understood their contributions. Some quotes were edited in this process to avoid harming relationships between organisations seeking to work collaboratively on mortality estimation, without substantively changing the quotes’ meanings.

## 3. Results

### 3.1. Mortality measurement initiatives and actors

Standardized Monitoring and Assessment of Relief and Transitions (SMART) surveys have been used by nutrition sector actors to collect information about crisis-attributable mortality for decades in Somalia, CAR and Cox’s Bazar (online Appendix B). In ‘full’ SMART surveys, anthropometry, mortality, and other variables are collected from a representative population sample, using standardised methods. They are primarily used to orient operational decision-making and may be done at a crisis-wide, district-or project-level scale. Mortality questionnaire modules may be omitted if mortality is assumed to be under control, other sources of mortality data are available, or when surveys must be implemented rapidly, in which case nutrition is prioritised.

Although SMART is a global initiative, the main actors involved in SMART differed substantially across settings. Somalia was among the first countries where SMART tools were piloted in 2005 (SMART, n.d.). Piloting coincided with establishment of the IPC in 2004, which arose from a perceived need for an evidence-based prioritisation framework to allocate resources across and within crises (IPC, n.d.). In Somalia, a specialized unit of the UN Food and Agriculture Organization (FAO) not found in other countries (the Food Security and Nutrition Analysis Unit, FSNAU) carries out most SMART surveys through a network of experienced Somali field researchers supported remotely. The FSNAU’s future vision is to be managed by Somali governing authorities, but in reality collaborations with government are limited (Maxwell and Hailey, 2021) and the unit’s website emphasises its independence: “[FSNAU’s] independence ensures neutral and objective analysis which should not be compromised in such a complex political and humanitarian environment” (FSNAU, 2010). More than 300 district-level SMART surveys have collected mortality data in Somalia over the past two decades, compared to four nation-wide surveys in CAR (Appendix B). In CAR, surveys are organized and funded by UNICEF (through donors), and implemented by the government’s statistics institute, the Institut Centrafricain des Statistiques et des Études Économiques et Sociales (ICASEES). ICASEES often draws on staff and students at the University of Bangui to carry out data collection for both SMART and Multiple Indicator Cluster surveys (MICS), which also measure mortality albeit over timespans of five years and with demographic methods not designed for the timelines of crises. In Cox’s Bazar, the only full surveys have been implemented at district-level by the NGO, Action Contre le Faim, which hosts the global SMART Initiative and had been conducting surveys among the host population since 2011.

An abridged mortality questionnaire has also recently been incorporated into annual nation-wide Multi-Sectoral Need Assessment (MSNA) surveys in Somalia and CAR by the NGO, IMPACT (REACH, 2025a, 2025b). IMPACT works through ACTED, an NGO with a large operational presence that uses local teams to collect data across the country. Commissioned by OCHA to inform annual humanitarian response planning, MSNAs are typically conducted in 15-20 crises per year, including Cox’s Bazar, but have not historically measured mortality. MSNAs are coordinated through OCHA at country level; health-related questions including mortality are agreed upon with the health cluster. IMPACT then incorporates available primary and secondary mortality and other data into situation analyses using the ‘public health information domains in crisis’ framework (Checchi et al., 2017).

Independent mortality surveys, which usually use standard cluster survey sampling methods, have also been undertaken by NGOs, foreign technical agencies and universities in all settings for decades. Often conducted in new or neglected phases of crises, they have been used not only to guide programmes but also to emphasize the impacts and politically-sensitive drivers of crises, including actions of governments and UN institutions. MSF’s 2017 survey in Cox’s Bazar documented the impact of violence experienced by Rohingya people in Myanmar to advocate against refoulement by the Bangladesh government (MSF, 2017). A 2022 survey in CAR by the Congolese NGO, Rebuild Hope for Africa (RHA) with foreign academics and Central African experts, was conducted to draw attention to mortality caused by the government’s use of Russian Wagner mercenaries and challenge the UN’s reliance on government-produced demographic data for humanitarian planning and development monitoring. The RHA team described how “people were seriously suspecting that the Wagner and the CAR government were carrying out massacres within populations. But there was no one who had […] proven facts” (CR3) and “It may be that host governments and the allied United Nations that support them are not capable of producing reliable mortality estimates during conflicts” (Gang et al., 2023, p. 7).

In all settings, community-based surveillance (CBS) systems have been used to monitor the morbidity and mortality of populations at risk of high mortality outside of health facilities, including refugees and internally displaced persons (IDPs) (Caleo et al., 2012; e4c, 2023; Ratnayake et al., 2020; Van Boetzelaer et al., 2020). Unlike surveys, which measure mortality retrospectively, surveillance systems produce mortality estimates prospectively (over time), requiring long-term, local capacity for ongoing data collection. UNHCR, which works in all settings, considers mortality monitoring to be part of both its health and protection mandates in refugee settings and routinely organizes partner organisations to contribute to continuous mortality surveillance and response (UNHCR, 2023). This way of working supports their aim of accountability to populations, as described by a UNHCR representative in CAR:

> “If even one death occurs in the community it is our responsibility to know what happened […] I call our partners every Monday to discuss the deaths and see what we can quickly do to improve […] it’s also our responsibility to bring the support to the family which has been affected by the death, so we have some SOPs [standard operating procedures] regarding protection aspects” (CR2).

Using community health workers to counsel, refer and tackle misinformation and stigma associated with certain kinds of deaths, such as stillbirths and early neonatal deaths or those due to COVID-19 was also seen as a benefit of introducing enhanced surveillance procedures for these mortality categories into the CBS system in Cox’s Bazar (Amsalu et al., 2022; WHO Bangladesh, 2021).

In Somalia, hybrid surveillance-survey methods have been used to monitor the evolution of mortality in sites hosting vulnerable and/or displaced populations. The Nutrition and Mortality Monitoring System (NMS) was set-up by the NGO Concern Worldwide and University College London in the UK, and was later expanded by e4c, and the Building Resilient Communities in Somalia (BRCiS) and Caafimaad Plus consortia. It has carried out repeated surveys within all households in purposively selected sentinel locations to produce mortality rates (e4c, 2023), while SIMAD has done so on a smaller scale among Mogadishu IDPs. Both of these initiatives have included verbal autopsy questionnaires to identify probable causes of death (Adam et al., 2025; Seal et al., 2021).

The large number of SMART surveys in Somalia has also enabled crisis-wide statistical modelling to estimate excess deaths by district-month and overall compared to a counterfactual baseline (FAO and FEWS-NET, 2013; UNICEF, 2025; Warsame et al., 2023):

> “the SMART Initiative has had beneficial impacts in the way that people tend to follow […] more or less, exactly the same protocol, and analyze their data using the same software. So that has helped to allow people to compare the data […and pull] together the data to come up with estimates of excess mortality” (SM2)

In Cox’s Bazar, modelling was also used to estimate COVID-19 deaths, incorporating data from the CBS system, one mortality survey, and other sources (Truelove et al., 2021).

### 3.2. Common political, operational and bureaucratic constraints and solutions

Actors across settings faced interconnected challenges that spanned financial, operational, political, and bureaucratic dimensions, each compounding the difficulty of obtaining and using mortality data for decision-making.

#### Financial constraints

Initial response to new crises brought additional financial resources for mortality measurement work in Somalia and Cox’s Bazar that were difficult to maintain as crises became chronic.

In Somalia, SMART survey work expanded during the 2017 and 2022 drought-triggered crises and donors that co-fund FSNAU also agreed to fund the NMS during these periods to provide additional data for rapid decision-making. Whereas SMART surveys in Somalia are typically done twice annually and take time to undergo validation and publication checks, the NMS’s shorter questionnaires and lightweight review and processing procedures enabled monthly reporting.

> “With say, an FSNAU [SMART] survey […] it’s very expensive and it’s a big operation, and there’s quite a lag time between deciding to do the survey and then getting the data. So I think the donors and others stakeholders were keen to see a system which was more flexible and adaptable, and nearer to real time data.” (SM2)

As drought responses tapered down, the NMS and statistical modelling efforts faced debilitating funding gaps, to the extent that they sometimes lost the capacity to function and thus identify further deteriorations signalling renewed crisis conditions. The time lag needed to restart both initiatives, while the 2022 drought emergency was escalating, meant that findings became available probably only after the emergency peaked (e4c, 2023).

In Cox’s Bazar, efforts invested at the beginning of the refugee influx to mobilise NGOs to enroll and train volunteers created a system that has been largely sustained eight years on, though partner funding gaps sometimes led to reduced reporting coverage, including in the lead-up to COVID-19 (UNHCR and WHO, 2020). Informants also raised concerns about the low levels of remuneration to volunteers.

In CAR, funding for SMART surveys has always been scarce and UNICEF reported being unable to fund any since 2022, potentially mirroring a global trend for chronic crises:

> “for many years SMART surveys were used and it was really useful to get all the mortality data from them, they were funded quite easily, even twice per year in many countries, but now […] it is more and more difficult to fund SMART surveys in countries when the crisis starts to be a bit too long.” (CR8)

#### Security and operability

As countries with areas controlled by non-state armed actors, Somalia and CAR had similar security and operability challenges. Survey work was hampered by large-scale flooding and poor road infrastructure, which was often worse in remote areas outside government control: “it just is a really hard place to have steady food supplies, steady drug supplies […] in the rebel held North, logistics and getting trucks through is really, really, really hard” (CR7). All nation-wide surveys in CAR had to exclude some populations because of a combination of insecurity, flooding or other logistical factors (see, for example, (Gang et al., 2023; MEPC and MSP, 2022; REACH, 2025b)); ICASEES-led surveys, being government-affiliated, were especially likely to exclude areas inaccessible to government.

There were strong disincentives for some kinds of actors to work in areas deemed insecure by UN peace-keeping missions supporting the government: “We had to hire groups of motorcycles so that we could go and assess in hidden parts. This is what the UN never, never do in their entire lives. And whoever within who tried would be fired for security [reasons]” (CR3). In Somalia, where the US military supports the government, agencies that receive funding from the US were also discouraged from working in areas controlled by al-Shabaab through application of bureaucratic controls to ensure aid cannot be diverted to them:

> “certain agencies were told to leave al-Shabaab areas at certain times, and other agencies perhaps chose to withdraw because of […] [US] OFAC [Office of Foreign Assets Control] regulations preventing agencies operating in areas controlled by what they had defined as a terrorist organization.” (SM2)

The NMS had to work mainly in areas under government control, but reasoned that collecting mortality information from populations displaced into these areas gave them an indirect understanding of patterns in areas held by al-Shabaab. With attacks against government targets happening across Somalia, some NGOs described data collection teams operating strict security protocols. To access IDP settlements, teams had to negotiate with armed security and settlement representatives frustrated by inadequate humanitarian assistance: “sometimes […] they were detaining our staff who were doing the data collection and [say]ing if you are not bringing anything, you cannot come back here” (SM4). In these situations, recruiting staff from local NGOs who had existing relationships with camp leaders was vital to smooth interactions.

In CAR, NGOs faced challenges engaging government actors to take action based on mortality findings in areas outside government control, such as when the International Rescue Committee (IRC)’s CBS programme reportedly identified hundreds of cases and deaths from suspected hepatitis E: “500 people died in that outbreak, and we could not get the Ministry of Health to go out […] and confirm [it]” (CR7). In Somalia, actors described the added complexity of working with so many autonomous state governments: “there’s lots of different controlling authorities now and so […] there are more layers of government to work with” (SM2).

#### Sensitivities and censorship

Mortality measurement was considered politically sensitive and subject to government influence in Bangladesh and especially CAR.

The RHA survey’s story illustrates the far-reaching effects of censorship. Following reports of increased insecurity, some members of IRC’s surveillance system proposed conducting a nation-wide survey. IRC leadership reportedly declined because it would be too politically risky, but MSF agreed and worked with the Ministry of Health and ICASEES to plan the survey and acquire approval from the government-appointed ethics committee, housed jointly within the Pasteur Institute in CAR and the University of Bangui. MSF and partners conducted the survey in one prefecture in 2020 (Robinson et al., 2021), but had to suspend work because of the COVID-19 pandemic. When they attempted to restart work two years later with a new ethics application, they received no answer from the committee for six months. They reportedly interpreted this silence as a rejection due to political interference or the fears of committee members and authorities that they could personally become targets of the Wagner group who, during the pandemic years, had become close to the government. MSF cancelled the survey because “they knew that if they go out and document what these Wagner soldiers have been doing [… they could] get kicked out of the country” (CR7). MSF handed the survey back to individuals involved in the original IRC project who formed a new team under RHA. As described by one team member, they were ultimately able to implement the survey through negotiations with local authorities in each area and the cooperation of a wide social network of people in UN agencies, NGOs and local authorities: “It was so flattering how these people wanted to support us. They essentially didn’t care what their bosses thought and would help us out,” for example, by securing plane tickets or writing protective letters for police.

Others have encountered bureaucratic challenges. Even with a lack of funding through UNICEF to conduct nation-wide SMART surveys in 2023 and 2024, NGOs with access to small scale funding struggled to get approval from local authorities for any surveys in 2024 because the government “requests to lead and implement all analysis processes […] This is a political issue. Just having so many NGOs in the country, it makes it difficult to harmonise information” (CR8). The only actor who has managed to implement mortality measurement in CAR recently, IMPACT, relied on high-level negotiation by UN-OCHA and the health cluster to gain approval for their MSNA work: “they [OCHA] talk on our behalf […] make it clear that we’re not trying to do anything fishy […] We’re not trying to make human rights statements […] We’re trying to inform […] decisions” (CR5).

We also found examples of censorship in mortality reporting. One informant described government actors “covering up” a methodological mistake that potentially showed reduced mortality in the 2018 SMART survey report, after the issue was raised in a nutrition cluster meeting. In 2023, days after the Wagner group’s attempted coup in Moscow, the Minister of Health called a press conference to condemn the RHA study, calling it a “smear campaign” which blamed Wagner troops for mortality in CAR without evidence, driven by “geopolitical and geostrategic bias” (Ngrebada, 2023). He also attempted to have the study publication retracted. Two years on, censorship appears to persist. The RHA study is the only recently conducted survey excluded from IMPACT’s analysis of secondary mortality sources used to interpret their own mortality findings from the MSNA (REACH, 2025b), despite the RHA survey reporting the “highest measured nationwide mortality rate in the world” (Gang et al., 2023, p. 1), suggesting a ‘Goldilocks solution’ may have been reached here.

In Cox’s Bazar, we saw censorship around a reproductive age mortality study organized by the UN Population Fund (UNFPA) and the US Centers for Disease Control (CDC) to measure maternal and perinatal mortality during the first year of the crisis. Study findings were presented at the federal level but the maternal mortality ratio measured was reportedly too high for the full report to be released publicly. Only an executive summary (Handzel et al., 2019) was allowed to be circulated to humanitarian actors to inform the response, while the perinatal findings (Amsalu et al., 2022) were published several years later: “We conducted the analysis, [….] but […] at that time the Ministry of Health had concerns about releasing the data publicly and preferred to withhold publication […] because the mortality data was higher” (CB3).

Sensitivities around measuring mortality also extended to the Rohingya refugee community: surveillance teams reported needing to consistently reassure households about the intention behind collecting data and whom data would be shared with. These dynamics reflected a historical fear of government surveillance among Rohingya people (ACAPS, 2020) and threats to Rohingya CHWs’ employment from Camp in-Charge’s (CICs) affiliated with the Bangladesh government who insisted the former share individual decedent information directly, rather than in aggregate through UNHCR: “They [CICs] warned them [CHWs] that if you do not provide the information, I am going to stop your activity […] we have passed many hot talks with the government on that thing” (CB7).

Mortality measurement appeared less sensitive in Somalia, where government approval for mortality study protocols and results validation happens via the nutrition sector’s Assessment Information Management (AIM) Working Group, which is co-chaired by the Ministry of Health and humanitarian partners, and rarely obstructs work: “the Ministry of Health […] might send you back, try to change some things. But once you get their blessing and go ahead, then, things become a bit smoother later on” (SM1). Government actors also made supportive statements at the launch of two SIMAD and LSHTM mortality studies presenting high excess mortality (Adam et al., 2025; UNICEF, 2025).

#### Coordination and data sharing

Across settings, UN coordination systems provided a backbone for regular mortality measurement and new initiatives, but systematic problems with data sharing created inefficiencies and led to potential duplication of efforts.

The first year of the response in Cox’s Bazar saw multiple, parallel mortality measurement activities, with UN inter-agency dynamics complicating coordination. Activities included MSF’s survey in November 2017, and ACF’s SMART surveys run every six-months from Oct 2017 until malnutrition rates had improved and death rates fell below emergency thresholds for South Asia (ACF, 2018). They also included efforts to implement continuous CBS reporting. Prior to the refugee influx, UNFPA had been operating a maternal and perinatal death surveillance and response (MPDSR) programme in Bangladesh which reviews all maternal, stillbirth and neonatal deaths, and, with CDC assistance, attempted to extend it to populations in the Rohingya camps as soon as refugees began arriving. UNHCR and WHO guidelines for surveillance in crisis settings both recommend delaying intensive MPDSR activities until lifesaving health services and a crude mortality surveillance system are in place (UNHCR, 2023; WHO, 2021). UNFPA and CDC nevertheless argued they could, from Sep 2017, train community health workers to complete both a weekly general mortality report and audits of maternal and perinatal deaths for the first time in a refugee setting. They agreed to adopt UNHCR’s simpler deaths reporting tools to streamline procedures. By mid-2019, UNHCR had developed a terms of reference for all NGO partners across camps to allocate community health workers to the surveillance initiative, with additional reporting for maternal and perinatal deaths and suspected disease outbreaks (UNHCR and WHO, 2020; Van Boetzelaer et al., 2020). Several informants emphasized the challenging discussions across working groups that were required:

> “that was also very challenging during the initial period to make all the partners realize the importance of comprehensive mortality surveillance so that we do not stick with any specific [categories of] deaths […] but we have successfully overcome that challenge after too many meetings” (CB7).

In Somalia, FSNAU’s SMART work over two decades has provided core evidence to classify multiple emergencies through the IPC and its continuity of surveillance between emergencies has helped maintain infrastructures that others have used to develop innovations. LSHTM-SIMAD modelling studies are entirely reliant on SMART data, and improved with the expansion of SMART surveys (UNICEF, 2025). The NMS emerged from a 2016 evaluation of the nutrition impacts of Concern’s cash-transfer intervention for Mogadishu-area IDPs and benefitted from the nutrition sector’s study approval and data validation systems. When the IPC projected a risk of famine in February 2017, members of the NMS approached the UK’s FCDO to continue funding the surveillance system, reframing its possibilities as a way “[t]o provide sentinel site data during the evolving emergency” and sharing monthly reports with responding agencies through the nutrition and health clusters (Seal et al., 2021, p. e1288). The system was reinstated again for the 2022 emergency, this time through two existing NGO consortia, BRCiS and Caafimaad Plus, which enabled the system to expand to a network of seven IDP sites across the country with “many of the heavyweight health and nutrition actors being part of the system” (SM1) and benefit from joint policy influence through the consortium’s different donors. As donors became a regular audience, they began requesting follow-up actions from cluster leads based on the findings. One observer described: “[it] really worked […] people are looking at the data and know what needs to happen” (SM1).

However, alongside these innovations by CDC, academics, NGOs, and donors, lack of collaboration by some UN agencies held back data sharing. SMART survey datasets and reports were not routinely shared in a predictable and accountable manner in any setting. Accessing and curating UN-held datasets for statistical modelling was perceived by the LSHTM-SIMAD team as the main challenge, a problem worsened by an FSNAU decision to stop publishing SMART datasets between the 2011 and 2017 emergencies: “[SMART survey] data is very fragmented all over the place. [… It takes] quite a lot of time negotiating access and then putting the data back together and then cleaning it” (SM3).

UN preferences also added complexity to scaling approaches by partners. During the 2022-23 drought, WHO declined a SIMAD proposal to use WHO funding for verbal autopsy research in IDP sites to join the NMS system, preferring to have more direct control over the funding (Adam et al., 2025). SIMAD collected demographic data in separate sites in parallel to the NMS, sending regular reports to WHO to inform outreach efforts, however many people involved felt an opportunity was missed to coordinate activities and reporting to strengthen local systems.

Finally, many informants described how nutrition and health actors were sometimes unaware of each other’s mortality measurement plans. This may explain why, in 2022, the health clusters in CAR and Somalia requested IMPACT incorporate mortality measurement in future annual MSNAs – despite all the mortality measurement work being done at this time by UNICEF/ICASEES and RHA in CAR and e4c, SIMAD, FSNAU, and LSHTM in Somalia – “because they didn’t have access to all the data already being collected” (2025 stakeholder meeting participant).

### 3.3. Impact of mortality measurements on humanitarian decision-making

In Cox’s Bazar, the COVID-19 pandemic exposed some limitations of UNHCR’s CBS system, prompting international debate about how so few deaths could be reported from a context with such high transmission risks (UNHCR and WHO, 2020). One investigation by the International Organisation for Migration suggested the Rohingya community were hiding deaths and identified mosque committees as a more reliable source about deaths and their circumstances than the official CBS (ACAPS, 2020). A Johns Hopkins/MSF model based on this data estimated only 1 in 62 deaths were reported during March to October 2020 (Truelove et al., 2021). A UN actor recalled an impact of this investigation was to supplement CHW reporting with grave-counting, and though limitations of this method were acknowledged, it resolved the debate for some people: “we started to look into how many people get buried in the graveyards […] from our perspective, we felt confident that there is not, you know, 100% more [deaths]” (CB1). The legacy of instituting maternal and perinatal deaths measurement early in the crisis appeared more successful. As of 2024, community-based sources provided the information on 91% of deaths in women of reproductive age known to the health sector, and annual MPDSR analyses, no longer censored, provide information to drive interventions that address an often overlooked area of mortality in crises (Health Sector, 2024).

In CAR, informants cited few examples where mortality data led to public health actions or funding changes, possibly due to stifled discussion in coordination meetings and opaque UN processes. One informant noted a lack of transparency in OCHA’s use of MSNA evidence: “I know they [OCHA] took it [the MSNA analysis] and accepted it and acknowledged it […] but past that, what decisions did they actually make? […] I’m not sure.” To improve transparency and increase use of mortality estimates by decision-makers, IMPACT began using existing systems to approve protocols and validate results in Somalia and CAR in 2024. A stakeholder meeting participant emphasised the need for cultural change in how OCHA shares and uses mortality data. Although OCHA’s Joint and Intersectoral Analysis Framework calls for using mortality data, it is rarely featured in their information systems or analytic products. Even when available in CAR, OCHA staff reportedly use mortality data “furtively” due to political sensitivities.

The way SMART reports are written may also influence decisions. With most space dedicated to technical aspects of nutrition measurement, mortality findings sometimes appear buried. Since the civil war began in CAR, mortality has been discussed only once in any report’s conclusions or recommendations, in 2014, simply as having an “upward trend” (ICASEES, 2015, p. 12), despite all surveys measuring mortality above the emergency threshold in several prefectures. Many recommendations also appear unrealistic and pro-government as in this one from 2022: “Encourage the government to strengthen national territory security in order to boost food production activities within the country’s different communities” (MEPC and MSP, 2022, p. 19). This language differs markedly from that in a comparable report from Somalia’s more independent NMS in 2023: “the outlook over the next few months remains concerning and emergency response capacity should be retained. The military action against Al-Shabaab by government forces, local clan militia, and international forces continues […] raising the possibility of further population displacement” (e4c, 2024, p. 22).

In CAR, while donors reportedly “believed” the RHA survey findings, global aid cuts prevented any donor from pursuing new actions. The independent RHA survey and Somalia statistical studies reportedly had some high-level influence, however; authors were told by the UN’s global statistics office that their data had been incorporated into modelled demographic estimates for CAR and Somalia in 2024 – the first time for a non-governmental source (UN Population Division, n.d.).

Famines in Somalia have a history of prompting large scale action, and mortality estimation has been intimately connected to this logic. As shared in a mortality meeting in 2024, the shocking levels of death recorded in a CDC study in Baidoa in 1992 (Moore et al., 1993) provided evidence for the US’s invasion the next year. Aid funding increased substantially following a declaration of famine in 2011 (Maxwell et al., 2023). The LSHTM model, however, suggested that a high proportion of people had died before the famine threshold was crossed (FAO and FEWS-NET, 2013), and advocacy work around this study is widely credited with more timely responses to crises in 2017 and 2022 which averted more deaths (Maxwell et al., 2023): “All the humanitarians said that their positioning had changed […] they specifically cited those previous studies as being an impetus for scaling up early” (SM3). In a stakeholder meeting, the NMS was also discussed as influencing decisions through providing evidence for IPC classifications in 2022 and by regularly demonstrating the differences between “supposed needs and actual needs on the ground”. A donor argued: “The NMS had a real-time impact […] if we’re going to save lives, that’s what we need to fund. And it isn’t expensive – these methods aren’t the big expenses when thinking about aid delivery.”

Finally, in both CAR and Somalia, informants highlighted the need for improved approaches to develop, coordinate and scale-up mortality measurement to better inform decision-making:

> “I think what UNHCR is doing [with their CBS system], and MSF also is working […] to promote the retrospective mortality survey [at smaller scales…] it could be good to introduce partners to ways to produce mortality data in other ways than formal [nation-wide SMART] surveys” (CR8);

> “if you look at that web of what’s being carried out at the moment, none of [the mortality measurement initiatives] are predictable, reliable or sustained. And if you look at the frequency of near famine or drought situations or major disease outbreaks, I think it creates a major challenge for us, and there must be a more cost effective […] more predictable solution” (SM6).

## 4. Discussion

By tracing the diverse mortality measurement work of actors across Somalia, CAR and Cox’s Bazar we have identified several common challenges to systematic data collection and use in humanitarian decision-making. Insecurity, disincentives and denial of permissions for actors to measure mortality in areas outside government control routinely obstructs coverage in CAR and Somalia. Combined with the effects of censorship on actors whose mortality measurements could draw attention to the state’s role in causing or maintaining high mortality in CAR, these coverage gaps likely have cumulative effects which limit actors’ understanding of vulnerability and interventions that might be needed. In all settings, the onset of new crises (food emergencies, refugee influxes, COVID-19) stimulated financial resources and innovations for mortality measurement, though they were not always sustained as crises resolved or became chronic. UN coordination systems provided a backbone for regular mortality measurement work that new initiatives built from, but problems with data sharing created inefficiencies and led to potential duplication of efforts.

Our analysis was limited to protracted crises with established humanitarian responses and government-humanitarian relationships; the ways of working we identified might not be possible in more recent crises, where logistics, access and censorship may incur bigger challenges. We did not manage to access representatives of crisis-affected governments, opposition groups or communities, most of our interviews were conducted remotely and we achieved low sample sizes; these issues limited our understanding of the use of mortality data for decision-making. Nevertheless, we comprehensively mapped mortality measurement activities across contexts and examined how actors interact with funders and systems of bureaucracy.

Our political economy approach highlighted how actors’ decisions on methodological approaches depended on actors’ own institutional histories, priorities and ways of working. They built on opportunities that emerged within specific crisis setting contexts, such as the need for a flexible sentinel surveillance system like the NMS in Somalia which could expand during emergencies and by-pass UN bureaucracies to report regularly to a wide range of actors. UNFPA and CDC’s proposal to integrate maternal and perinatal death surveillance in the first month of programming in Cox’s Bazar had never previously been done in a refugee response and was possible because of the organisations’ existing work in this area elsewhere in Bangladesh. Innovations also emerged out of global analyses of challenges across crises, such as the need of OCHA and the health clusters for mortality information, which enabled integration of mortality measurement into the annual MSNA to emerge as a solution.

Our findings align with previous research on organisational behaviours in mortality measurement. Data sharing was a major challenge for FSNAU in Somalia and IPC actors elsewhere in 2017, often due to organisational competition and fear of methodological criticism (Maxwell and Hailey, 2021). In CAR, we observed various forms of censorship, including overt government interference – such as suppressing reports and analyses, and threatening individuals – like in South Sudan, Sudan and Myanmar (Farley et al., 2024; Maxwell and Hailey, 2021; McPherson et al., 2024). Quieter bureaucratic controls, such as MSF’s ignored ethics application in CAR, has also been experienced by members of our team attempting to do mortality work in other settings, and stifles evidence production (Storeng et al., 2019). While building government capacity is a key UN mandate and a form of localisation (Healy et al., 2020), in contexts where governments impede humanitarian work, collaboration with civil society willing to work without permissions seems ethical and legitimate. We also noted donor-driven censorship to defend investments in maternal mortality in Cox’s Bazar. Contrary to a 2007 analysis that identified NGOs as primary producers of SMART surveys, (Degomme and Guha-Sapir, 2007), in Somalia and CAR, UN and government dominate data collection, likely due to institutional histories and government control over NGOs in CAR.

As governments and humanitarian organisations adapt to major cuts in global humanitarian aid, reliable mortality data will become more crucial for prioritizing resources. It will be important to leverage different partners’ access to various populations to avoid focusing only on easily accessible areas. While crisis-affected governments may take on increasing ownership of certain areas of mortality measurement work, the sensitive nature of mortality data means independent actors and funding remain essential to ensure data are collected, prevent censorship and build capacity of diverse stakeholders. Consortia working under a technical coordinating agency may also help mitigate reputational risks in sensitive contexts. The NMS’s consortium approach aligns with current aid sector trends promoting sharing of power and complementarities between national and international NGOs. Academic involvement will remain vital to address technical questions and promote innovation.

## 5. End matter

### Author contributions

Jennifer Palmer: Writing – original draft, Writing – review and editing, Methodology, Investigation, Formal analysis, Visualisation, Conceptualisation, Supervision, Project administration, Funding. Mervat Alhaffar: Writing – original draft, Writing – review and editing, Investigation, Formal analysis, Project administration. Bashiru Garba: Writing – original draft, Writing – review and editing, Investigation, Formal analysis. Jihaan Hassan: Writing – review and editing, Investigation, Formal analysis. Ruwan Ratnayake: Writing – review and editing, Conceptualisation, Funding. Andrew Seal: Writing – review and editing, Conceptualisation, Funding. Mohamed Jelle: Writing – review and editing, Conceptualisation, Funding. Francesco Checchi: Writing – review and editing, Conceptualisation, Project administration, Funding.

## Acknowledgements

We would like to express our gratitude to the 23 informants who generously volunteered their time and insights to contribute to our research study and Daud Ibrahim Sheikh in Somalia’s Ministry of Health for introducing us to some informants. We also thank other members of the Mortality Estimation Systems Innovation Partnership study team and participants in three global stakeholders’ workshops who provided feedback on early findings.

## Funding source

This study was funded by the United Kingdom Humanitarian Innovation Hub (UKHIH, no grant number assigned) and its donor, the UK Foreign Commonwealth and Development Office (FCDO). The FCDO had no role in the study’s design, implementation, analysis, or dissemination of results, and had no role in the writing of this article. UKHIH provided advice on study design and assisted with dissemination of results to key humanitarian stakeholders but played no role in study implementation, analysis or article writing.

## Declaration of competing interest

We declare no competing interests.

## Generative AI declaration

During the preparation of this work the authors used perplexity.ai to simplify language in some paragraphs. After using this tool, the authors reviewed and edited the content as needed and take full responsibility for the content of the publication.

## Data availability

The data that has been used is confidential.

## 7.3 Appendices

Appendix A. Consolidated Criteria for Reporting Qualitative Research (COREQ) checklist

Appendix B. List of mortality data collection exercises, by case study context 1991-1992:

## Notes

### Competing Interest Statement

The authors have declared no competing interest.

### Author Declarations

Research Ethics Review Committees of the London School of Hygiene and Tropical Medicine in the United Kingdom (ref: 31136) and SIMAD University in Somalia (ref: 2024/SU-IRB/FMHS/P0012) gave ethical approval for this work.

