## Appendix A for "The politics and challenges of measuring mortality for humanitarian decision-making in Somalia, the Central African Republic and Bangladesh"

**Consolidated criteria for reporting qualitative studies (COREQ): 32-item checklist**

Framework from: Tong A, Sainsbury P, Craig J. Consolidated criteria for reporting qualitative research (COREQ): a 32-item checklist for interviews and focus groups. *International Journal for Quality in Health Care*. 2007. Volume 19, Number 6: pp. 349 – 357

| **No. Item** | **Guide questions/description** | **Response** |
| --- | --- | --- |
| **Domain 1: Research team and reﬂexivity** | | |
| *Personal Characteristics* |  |  |
| 1. Interviewer/facilitator | Which author/s conducted the interview or focus group? | Interviews were conducted by JP in CAR case study, MA in Cox’s Bazaar case study and BG & JH Somalia. JP conducted meeting observations and supervised data collection across all settings. |
| 2. Credentials | What were the researcher’s credentials? E.g. PhD, MD | JP, BG hold a PhD, MA is a PhD candidate |
| 3. Occupation | What was their occupation at the time of the study? | Both JP and BG are working as associate professors, while MA is a PhD student and research fellow |
| 4. Gender | Was the researcher male or female? | Both male and female |
| 5. Experience and training | What experience or training did the researcher have? | JP, MA and BG have established experiences in conducting qualitative research |
| *Relationship with participants* |  |  |
| 6. Relationship established | Was a relationship established prior to study commencement? | Several members of our wider research team had previously worked on mortality measurement in Somalia and knew relevant actors who participated in interviews or connected us to others. At the beginning of the study we participated in global mortality measurement meetings/events to meet some stakeholders who later participated in interviews about CAR, Cox’s Bazar and Somalia and introduced us to others. Members of our wider author/research team also provided observations based on their prior and ongoing work in Somalia. |
| 7. Participant knowledge of the interviewer | What did the participants know about the researcher? e.g. personal goals, reasons for doing the research | Information sheet that contained all the research objectives were shared prior to the interviews. |
| 8. Interviewer characteristics | What characteristics were reported about the interviewer/facilitator? e.g. Bias, assumptions, reasons and interests in the research topic | The interviewers’ background and how the interviews were conducted were reported in the manuscript. |
| **Domain 2: study design** | | |
| *Theoretical framework* |  |  |
| 9. Methodological orientation and Theory | What methodological orientation was stated to underpin the study? e.g. grounded theory, discourse analysis, ethnography, phenomenology, content analysis | Data collection and analysis was iterative, based on meetings, document review and interviews. Data was analysed manually using thematic analysis and informed by applied political economy theories. JP has experience of doing ethnography in policy analyses, which informed several aspects of the data collection and analysis (ways of reading documents, observing and noting meetings, iterative analysis through internal and external presentations and repeat interactions with informants). |
| *Participant selection* |  |  |
| 10. Sampling | How were participants selected? e.g., purposive, convenience, consecutive, snowball | Purposive and snowball sampling were used to identify informants based on their involvement in at least one mortality measurement initiative in each of the three contexts. For informants with multiple affiliations, we categorised them by the type of institution they were in when working on the main mortality initiative they spoke about in the interview. |
| 11. Method of approach | How were participants approached? e.g. face-to-face, telephone, mail, email | Emails were used to introduce the study, obtain consent and organize interviews. Some participants were initially approached in meetings. Interviews about CAR and Cox’s Bazaar were conducted remotely from the United Kingdom and a combination of in-person and remote interviews in Somalia. Only 2 people provided written responses to emailed questions only. |
| 12. Sample size | How many participants were in the study? | 23 key informants |
| 13. Non-participation | How many people refused to participate or dropped out? Reasons? | We did not receive responses from around one-third of people we approached for an interview, including from people who mentioned they would need to request permission from their organisational hierarchies because of the sensitivity of mortality measurement in the context. |
| *Setting* |  |  |
| 14. Setting of data collection | Where was the data collected? e.g. home, clinic, workplace | Private spaces at both home and workplace |
| 15. Presence of non-participants | Was anyone else present besides the participants and researchers? | No one else present during the interviews |
| 16. Description of sample | What are the important characteristics of the sample? e.g. demographic data, date | 70% of interviewees were men (100% in Somalia). Data collection took place between May 2024 and March 2025. |
| *Data collection* |  |  |
| 17. Interview guide | Were questions, prompts, guides provided by the authors? Was it pilot tested? | A loose topic guide was developed. Before each interview, relevant information about informants’ organisations and the mortality measurement initiative they had experience of (from websites, documents, prior interviews and meeting observations) were reviewed to identify priority and additional questions to focus the discussion. |
| 18. Repeat interviews | Were repeat interviews carried out? If yes, how many? | We had multiple interactions with around one third of informants that consisted of a main interview with follow-up discussions to collect more information via email or during global mortality measurement meetings. |
| 19. Audio/visual recording | Did the research use audio or visual recording to collect the data? | Remote interviews were conducted via a mix of video and audio calls and audio recording was used with participants consent. We also audio recorded two of the stakeholder meetings we observed and took detailed notes in the remainder. We used Otter.ai software to produce transcripts in real-time. Authors simultaneously typed detailed notes and listened to audio files afterward to correct inaccuracies in the transcripts.  Researchers created their own summaries of key information they learned after each interview, rather than relying on summaries automatically-generated by the software. |
| 20. Field notes | Were ﬁeld notes made during and/or after the interview or focus group? | Yes |
| 21. Duration | What was the duration of the interviews or focus group? | Interviews lasted between 30-120 minutes |
| 22. Data saturation | Was data saturation discussed? | Yes |
| 23. Transcripts returned | Were transcripts returned to participants for comment and/or correction? | No |
| **Domain 3: analysis and ﬁndings** | | |
| *Data analysis* |  |  |
| 24. Number of data coders | How many data coders coded the data? | JP, MA, BG & JH all collected and analysed the data |
| 25. Description of the coding tree | Did authors provide a description of the coding tree? | A detailed, shared coding tree was not developed – instead we kept working papers using a shared sub-headings template to organise data from all sources about major topics and themes. Analyses evolved very iteratively through the process of preparing for and debriefing after frequent internal and public/stakeholder presentations to identify gaps in our knowledge and refine the ‘story’. |
| 26. Derivation of themes | Were themes identiﬁed in advance or derived from the data? | A mix of both |
| 27. Software | What software, if applicable, was used to manage the data? | Data was analysed manually for each case study |
| 28. Participant checking | Did participants provide feedback on the ﬁndings? | Yes, we shared a copy of the findings with participants for validation and ask their permissions to include quotes. Individuals at stakeholder meetings also provided feedback on our emerging analyses. |
| *Reporting* |  |  |
| 29. Quotations presented | Were participant quotations presented to illustrate the themes/ﬁndings? Was each quotation identiﬁed? e.g. participant number | Yes with participants ID which remains anonymous |
| 30. Data and ﬁndings consistent | Was there consistency between the data presented and the ﬁndings? | Yes |
| 31. Clarity of major themes | Were major themes clearly presented in the ﬁndings? | Yes |
| 32. Clarity of minor themes | Is there a description of diverse cases or discussion of minor themes? | Our analysis was a three-way comparison. When discussing each theme we consistently considered how it applied in each of the settings, as well as to each key organisational actor within settings. |
