## Appendix B for "The politics and challenges of measuring mortality for humanitarian decision-making in Somalia, the Central African Republic and Bangladesh"

Appendix B. List of mortality data collection exercises, by case study context

*Table B1. Mortality measurement activities across case studies with lead institutions and dates of data collection in parentheses*

|  | **Central African Republic** | **Somalia** | **Cox’s Bazar** |
| --- | --- | --- | --- |
| **MICS and DHS**^1^ | MICS (UNICEF/ICASEES in 2000, 2006, 2010, 2018/19)^1^, DHS (1994/5)^2^ | MICS (UNICEF/Ministries of Planning in 1996/7, 2000, 2006, 2011 in Somaliland & Puntland)^1^ | - |
| **SMART surveys** | 4 nation-wide surveys (UNICEF/ICASEES in 2012, 2014, 2018, 2022)^3–5^ | >300 sub-national surveys (FSNAU, NGOs over ~2 decades)^6^ | 4 multi-camp surveys (ACF in 2017, 2018, 2020)^7,8^ |
| **Multi-sectoral needs assessments** | Mortality measured annually (IMPACT in 2023, 2024)^9^ | Mortality measured annually (IMPACT in 2023, 2024)^10^ | - |
| **Other mortality surveys** | NGO/academic surveys (UNICEF 2009, MSF 2012, MSF 2020, RHA 2022)^11–14^ | NGO/academic surveys (US CDC 1993, CDC 2011 (Dadaab), LSHTM 2021)^15–17^ | NGO/technical agency surveys (MSF 2017, IOM 2020)^18,19^ |
| **Hybrid surveillance-survey methods** | - | Repeated surveys in vulnerable populations until mortality decreases (NMS UCL/Concern/e4c/BRCiS/Caafimaad consortia intermittently 2015-pres, SIMAD 2022-23)^20–25^ | - |
| **Community-based surveillance** | Weekly reporting (MSF 2010-11, IRC 2018-21, UNHCR 2023-pres)^26,27^ |  | Weekly reporting (UNHCR + 25 orgs 2017-pres)^28,29^ |
| **Crisis-wide statistical models** | - | During/after large-scale crises (LSHTM/partners 2010-12, 2014-18, 2022-24)^30–32^ | During COVID-19 (Hopkins/MSF 2020)^33^ |

Note: No DHS or MICS surveys have been conducted in Somalia for decades, apart from in Puntland. In Cox’s Bazar refugee camps, demographic monitoring is undertaken by humanitarian agencies rather than by government actors; nation-wide MICS surveys run by the Bangladesh Bureau of Statistics have excluded Rohingya refugees to date.

**Appendix B References**

9.	REACH. *Mortality Rates in Central African Republic: An Integrated Public Health Analysis*. Impact Initiatives; 2025.

28.	UNHCR, WHO. *Community Based Mortality Surveillance in Cox’s Bazar Rohingya Camps – 12 Month Brief (July 2020)*.; 2020.
